# Evaluating the Impact of NHS Strikes on Patient Flow through Emergency Departments

**DOI:** 10.1101/2024.09.03.24312252

**Authors:** Alex Garner, Quin Ashcroft, Dale Kirkwood, Vishnu Chandrabalan, Hedley Emsley, Suzanne M Mason, Nancy Preston, Jo Knight

**Affiliations:** Lancaster Medical School, Lancaster University; Lancashire Teaching Hospitals NHS Foundation Trust; School of Health and Related Research, The University of Sheffield; International Observatory on End of Life Care, Lancaster University

## Abstract

**Background:** Since December 2022, the NHS has experienced large-scale strikes over pay by staff. Strikes heavily impact elective care delivery. The NHS cancels approximately 12 million elective care appointments each year. One million appointments have been cancelled due to strikes between 2022 and 2024. During this time emergency care is prioritised, and in a recent opinion piece, the president of the Royal College of Emergency medicine claimed the Emergency Department ran ‘better than usual’. The aim of this paper was to investigate changes in patient flow into hospitals through the ED during the strike periods.

**Methodology:** Data from two different emergency departments (EDs) in the North West of England is analysed using Cox-regression to model time between patient arrival at the ED, and subsequent admission. Various systematic and patient-level factors are controlled for. The impact of different striking groups (nurses, junior doctors etc.) on patient time to admission is analysed.

**Results:** For the Type 1 ED, hazard ratios indicate that patients are admitted through the ED more quickly on strike days where any single group of staff were striking compared to non-strike days (HRs: 1.16-1.39, all p ≤ 0.003). This increased flow was only seen for consultant strikes in the smaller ED.

**Interpretation:** These findings for all strike types indicate that improved patient flow on strike days is likely due to the increased inpatient capacity from elective care postponement. This result may indicate that there is room for change in NHS hospital systems to improve turnaround time and reduce ED crowding.

**Key Messages:** *What is already known on this topic:* Exit block is a primary issue for NHS Emergency Departments (EDs), increasing patient time spent in ED. The president of the Royal College of Emergency Medicine claimed that NHS EDs run ‘better than usual’ during strikes. There is little quantitative evidence to support this claim.

*What this study adds:* This study identifies significant increases in flow of admitted patients into the hospital during the strikes, when accounting for differences in admission patterns.

*How this study might affect research, practice or policy:* The improvement in flow indicates that, although strike day service is unsustainable, elements of strike preparation can have a positive impact on the running of the emergency departments - demonstrating scope to improve patient flow through NHS EDs.

## Introduction

Since December 2022, the NHS has experienced large-scale strikes over pay and working conditions by junior doctors, consultants and other staff (1). In June 2023, nurse strikes in England ended due to low voter turnout. In April 2024, consultants agreed to end their strikes with a new pay deal (2). At the time of writing junior doctor pay deals are unsettled (3). By the end of 2023, the NHS estimated the costs of the strike amounted to around £1.5bn (4).

During strike periods, much routine care has been rescheduled and approximately one million appointments were cancelled between February 2022 and January 2024 (5,6). This contributes to the approximately 12 million elective care cancellations each year between 2021 and 2023 (7). Cancellations impact patient outcomes, with a survey by *Healthwatch* finding that 66% of people with cancelled care for any reason during 2023 said it had impacted their lives (8).

Emergency care provided by the NHS is protected and prioritised during strikes. Non-striking staff are often drafted in to cover the ED (6). The process by which patients are admitted to hospital through the ED typically does not change during strikes, only the mix of staff making the decisions.

Evidence from previous strikes has shown that strikes are associated with fewer emergency attendances, leading to less crowding in the emergency departments (9). ED crowding is generally associated with an increased risk of in-hospital mortality, longer times to treatment of patients with certain conditions and a higher probability of leaving the ED against medical advice or without being seen (10). Studies conducted under non-strike conditions suggest that the primary cause of ED crowding is exit block (or access block) (11,12). Exit block is defined as when ‘patients in the ED requiring inpatient care are unable to gain access to appropriate hospital beds within a reasonable time frame’ (13).

Studies indicate that healthcare worker strikes are correlated with lower patient waiting times in EDs globally (14). In an opinion piece, the President of the Royal College of Emergency Medicine, suggests that during some of the strikes ‘in the emergency department, everything works better than usual’ (15). Streamlined decision making during the strike periods is alluded to, while also suggesting wider hospital measures that make emergency department procedures more efficient during these times. One of these possible factors is more inpatient capacity due to the lack of elective activity in this period and consequent reduction of exit block (15).

In this paper, we aim to investigate these claims. We hypothesise increased patient flow in ED on strike days and seek to provide quantitative evidence for the NHS to support consideration of changes in ED practice. In this paper we analyse data on time spent in two emergency departments based in the North West of England. We use survival analysis methodology to model patients’ time from arrival in ED to admission controlling for other influential variables and evaluating the impact of the strikes.

## Methodology

### Data

The dataset structure is based on the NHS Emergency Care Data Set specification, a national specification for datasets from NHS emergency departments, set by NHS England and the Royal College of Emergency Medicine (16). The data covers attendances to two emergency departments in the North West of England between January 2022 and April 2024, giving time of arrival, time spent in ED, investigations performed and other demographic and diagnostic information. This data includes any attendance from patients who have not opted-out of their NHS data being used for research. We present our analysis according to the RECORD reporting guidelines.

Both EDs are operated by the same Trust. ED1 is a 24-hour, full-service ED with a major trauma service and averages around 50,000 attendances per year (since 2022), ED2 is an adults only minor injuries unit with limited hours and averages around 25,000 attendances per year (since 2022). People with major injuries arriving at ED2 are sent to ED1. The EDs are analysed separately, this paper focuses on the effects of the strikes at the major ED1 as results will be more generalisable to many other EDs. Full results for ED2 will be found in the supplementary materials and findings will be discussed in the main paper.

### PPIE Statement

ED staff were involved in the initial formulation of this project’s research question. No patients or members of the public were involved in this research.

### Outcome

The outcome in this analysis was time spent in ED for patients admitted to hospital directly from their ED attendance as an indicator for patient flow through the emergency department. Defined as the time between their arrival and departure from the ED. The departure time is defined as the time a patient is discharged or transferred from the emergency care attendance to a ward as defined by the NHS data dictionary (17). We use the subset of attendances that are transferred to a ward. From this point we refer to this as the “time in ED”.

“Time in ED” can be thought of as *time to event* data that can be used in a survival analysis. In this case, since the endpoint of interest – admission – is an outcome that is preferred more quickly, a higher hazard ratio is favourable as it reflects admitted patients spending less time in ED.

### Exploratory Testing for Strike Day Chanes

We used Kolmogorov-Smirnov two sample tests to assess the potential changes in number of admissions per day and proportion of attendances admitted to a ward during strike days and non-strike days over the study period.

### Kaplan Meier Exploratory Analysis

We investigated the proportion of patients that remained in ED over a continuous time period starting at their arrival at the emergency department. Kaplan-Meier plots were used to investigate empirical differences between factors that we stipulate could impact patient flow through the emergency department. Confidence intervals are calculated using Greenwood’s formula to estimate variance of the estimate at each time point (18).

### Cox-Proportional Hazards Modelling

Cox-Proportional Hazards models were then used to produce a semi-parametric regression model fitted to “ “time in ED” - fitted using the *lifelines* package in python (19). We created two models (for each of ED1 and ED2), each with five strike groups of interest to investigate the impact of the following strikes: *junior doctor strike, consultant strike, both junior doctor and consultant strike, nurse strike, ambulance strike*. All strike day categorisations can be found in Supplementary Table S1.

We also controlled for variables that could influence patient flow on strike days, or factors that may alter an individual’s time spent in ED, such as urgency of their presentation. Variables accounted for in the Cox-regression model are:

- ED Factors
  - Linear time effect
  - Seasonal effects – harmonic pair
  - Time of day – harmonic pair
  - ‘Heat’ – measure of number of patients in the ED scaled by urgency of those patients
- Patient Presentation Factors
  - Urgency of presentation
  - Referral service admitted to
- Patient Demographic Factors
  - Age
  - Ethnicity
  - Gender

We used the Cox model with standard errors calculated robustly via a bootstrapping method (20,21). Hazard ratios were calculated for each variable in the model, including the strike variable. The magnitude of the hazard ratios for junior doctor strikes and other strikes were tested at an α = 0.005 significance level - Bonferroni adjusted for the 5 strike types (compared to no strike) at each ED (10 tests). The proportional hazards assumption was assessed using log-log plots of empirical survival curves of the variables, and Schoenfeld residuals of the fitted Cox models (20).

## Results

There were 174,961 emergency attendances in the study period, 119,553 were to ED1. Of those, 49,165 (41%) resulted in an admission to the hospital. Observations where hospital admission status, referral service or urgency were missing in the data were not included, resulting in 44,229 admissions in the sample for investigation. Over the analysis period, we observed 61 separate strike days, the first was 15^th^ December 2022. There were 3,219 admissions during the strike days. Most of the strike day admissions (1,822 - 57%) occurred during junior doctor strikes. The patient demographics overall and specifically for strike days can be viewed in Table 1.

**Table 1:**
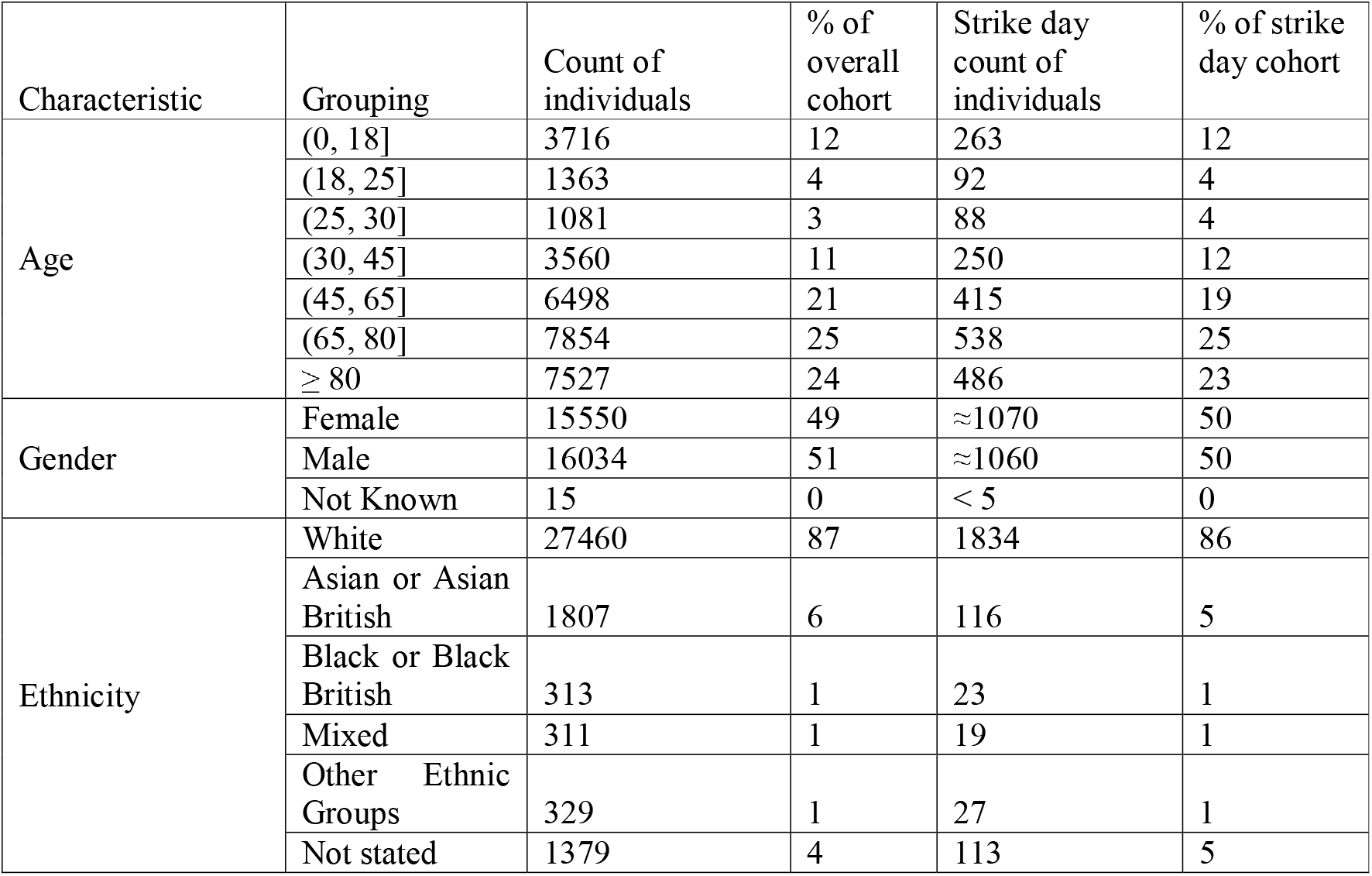
Demographic mix of patients overall and filtered for strike days.

| Characteristic | Grouping | Count of individuals | % of overall cohort | Strike day count of individuals | % of strike day cohort |
| --- | --- | --- | --- | --- | --- |
| Age | (0, 18] | 3716 | 12 | 263 | 12 |
|  | (18, 25] | 1363 | 4 | 92 | 4 |
|  | (25, 30] | 1081 | 3 | 88 | 4 |
|  | (30, 45] | 3560 | 11 | 250 | 12 |
|  | (45, 65] | 6498 | 21 | 415 | 19 |
|  | (65, 80] | 7854 | 25 | 538 | 25 |
|  | ≥ 80 | 7527 | 24 | 486 | 23 |
| Gender | Female | 15550 | 49 | ≈1070 | 50 |
|  | Male | 16034 | 51 | ≈1060 | 50 |
|  | Not Known | 15 | 0 | < 5 | 0 |
| Ethnicity | White | 27460 | 87 | 1834 | 86 |
|  | Asian or Asian British | 1807 | 6 | 116 | 5 |
|  | Black or Black British | 313 | 1 | 23 | 1 |
|  | Mixed | 311 | 1 | 19 | 1 |
|  | Other Ethnic Groups | 329 | 1 | 27 | 1 |
|  | Not stated | 1379 | 4 | 113 | 5 |

### Exploratory Testing for Strike Day Changes

We find that there is no significant difference between the number of attendances on strike days and non-strike days. We also find that there is no significant difference between proportion of attendances that are admitted on strike days and non-strike days.

### Average Outcomes

The subsequent analysis included only admitted patients, therefore “time in ED” always ends in admission of the patient in this analysis. We find that the median “time in ED” for the patients in the analysis was 18 hours 4 minutes (IQR: 8h 44m – 28h 47m). The median time before first contact with member of healthcare staff (*wait time*) is 2 hours 12 minutes. There is a high rate of admissions immediately before the four hour mark, which corresponds to patients being admitted immediately before the 4-hour target time set by the government (22). This surge in admissions before 4-hours skews the distribution of patients’ time spent in ED. Histograms of patients’ “time in ED” overall and for strike days are found in *Figures S1 and S2*. The median “time in ED” on a strike day is found to be 13 hours (IQR: 6h 58m – 23h 34m).

### Kaplan Meier Exploratory Analysis

We investigated differences in empirical “time in ED” curves between different categories, regardless of strike conditions. ***Error! Reference source not found***., demonstrates large variations in “time in ED” curves for the different services a patient is referred to. Paediatric had the fastest flow through the ED and General Medicine was the slowest, these different curves are likely to be in part the result of differing admission pathways between referral services and capacity in each of the different wards. The *Other* category contained any referral services with fewer than 500 admissions during the study period, the full list of services included in this category can be found in the supplementary materials. The Kaplan-Meier curves for the other explanatory variables are shown in the *Supplementary Figures S3 – S7*.

### Cox-Proportional Hazards Modelling Results

Coefficients and hazards ratios (of earlier admission) for the strike types are found in Table 2, the extended table for all variables is found in *Supplementary Table S2*. A visual description of these hazard ratios in Forest plot format is shown in Figure 2. Forrest plots for all variables are found in *Supplementary Figure S13*.

**Table 2:**
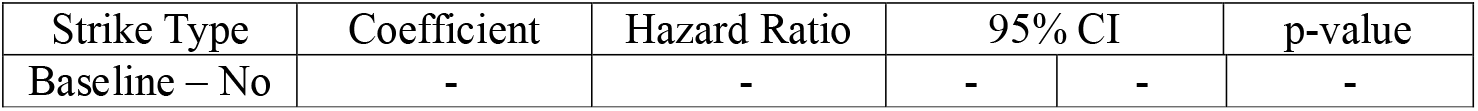
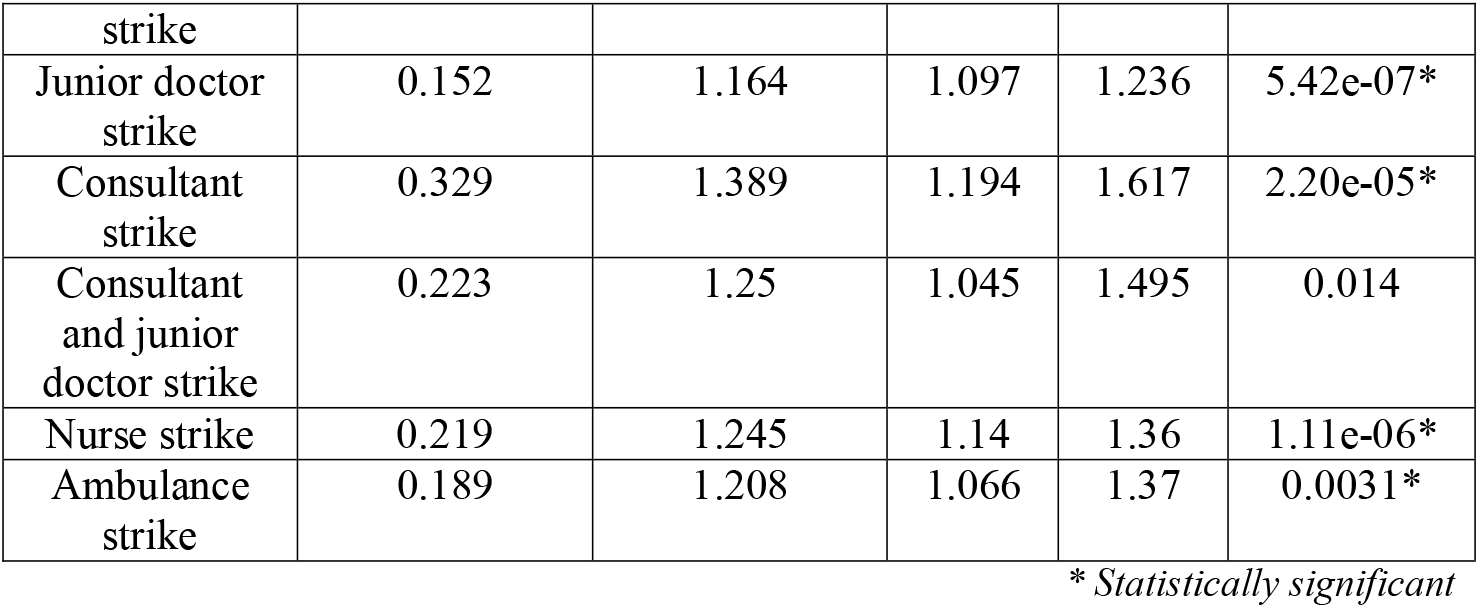
Coefficients and hazard ratios for the strike variables in the Cox-proportional hazards model. Here, higher hazard refers to a higher likelihood of being admitted into the hospital. Since we only investigate patients who were admitted, higher hazard is a positive outcome that means patients were more likely to be admitted more quickly.

The results suggest that all strikes have a similar effect on patient “time in ED”. The largest effect is for consultant strikes, with a hazard ratio of 1.39 suggesting that patients are admitted 39% faster on a consultant strike day compared to a non-strike day. The fitted survival curves from the models for all the strike types are shown in Figure 3. Since this only included patients who were eventually admitted to a ward, higher ‘hazard’ suggests that patients move through the emergency department into the hospital more quickly on strike days, leading to shorter “time in ED”.

The Schoenfeld residuals against transformed time tests indicated that five of the variables included in the model significantly deviated from proportional hazards. Inspected visually, these were minor deviations that were likely to be statistically significant due to large sample sizes. Results from these tests are found in the *Supplementary Figures S8-S12* considered further in the *Discussion* section. Results from ED2 are found in the supplementary materials under *ED2 Analysis*.

## Discussion

We provide the first quantitative evidence of the impact of the NHS strikes on patient flow into the hospital through the emergency department. Based on the hazard ratios we find a shorter time to admission from ED for admitted patients on all strike days, suggesting increased patient flow. Since this analysis only includes admitted patients, higher hazard ratio implies shorter “time in ED” on strike days than during non-strike days when controlling for ED and patient factors.

During strikes the hospitals were running unsustainable practices, and the authors do not suggest that healthcare worker strikes are positive overall. The issues of cancelled care and rising costs to the NHS have been raised in the introduction, cancelled elective care can lead to further ED attendances down the line. Furthermore, ED staff are experienced in making decisions whether to admit patients quickly and safely, and we do not investigate whether the decisions to admit in this dataset were appropriate. However, we identify that there was no increase in proportion of patients admitted during the strike periods. These results suggest that there may be scope to improve patient flow through emergency departments and reduce patient time spent in ED.

The shorter time in ED for admitted patients, indicating an increase in patient flow, is likely due to the hospital prioritising emergency care during the strike period. One of the causes could be additional capacity for patients admitted from the emergency department due to the reduction of elective activity in the hospital. This is compounded by rapid discharge of inpatients leading up to strike days. Specialists also often have additional capacity to deal with emergency patients (23). Reduction of elective care may also free up other hospital capacity, such as in the Intensive Care Unit. The apparent improved flow, suggested by shorter time in ED, is similar for all strike groups, suggesting differences in staff seniority between the strike types (although not measured directly in this analysis) is likely not a primary driver of this change.

The Royal College of Emergency Medicine reported in January 2024 that hospital bed occupancy rates were at the ‘unsafe’ level of 93% (24). Improved flow during strikes supports the hypothesis that exit block is the primary driver of ED crowding – indicating improved flow of patients out of hospital will likely improve waiting times in ED.

This finding agrees with some previous findings in the literature that imply a reduction in patient wait time during strike periods globally (25–27). However, findings are inconsistent and depend on the country’s healthcare system and the strike in question (14). Evidence from previous NHS strikes shows the negative impact on services and patient outcomes, but does not specifically investigate emergency departments (28,29).

An additional finding of the paper is the large variation in “time in ED” curves for the different referral services. Many of the differences between the different categories and the baseline of ‘orthopaedics’ in the fitted model are statistically significant overall (and vary between each other). The differences in survival curves between referral destinations demonstrated in Figure 1 are likely due to hospital flow structures. Flow and capacity issues that contribute to exit block vary between patient destinations.

**Figure 1:**
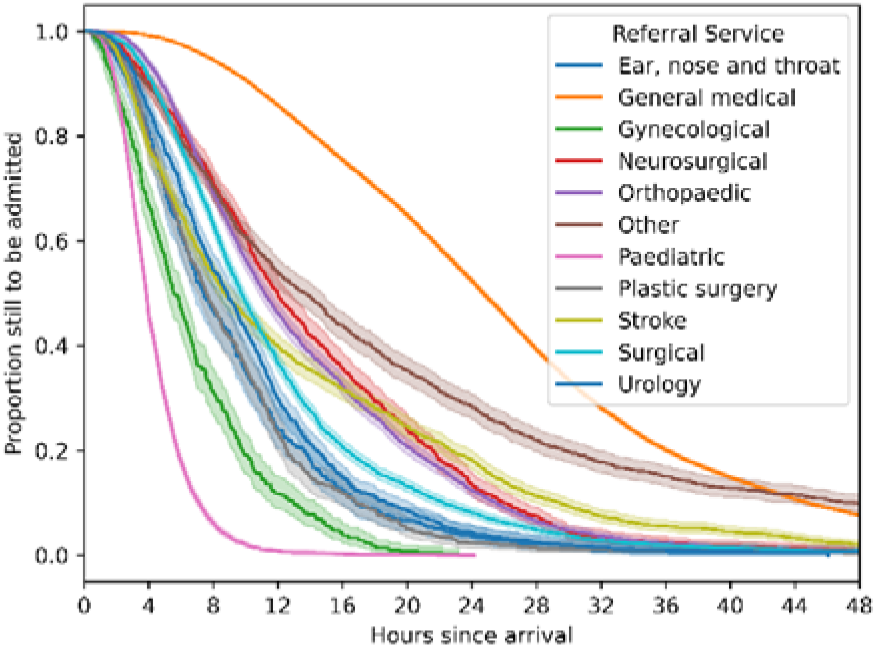
Kaplan-Meier curves of admitted patients’ “time in ED”, separated by referral service they are admitted to. The shaded area around each line represents the 95% confidence interval (CI) for the KM estimate at each point. These CI are very small for paediatrics and general medicine therefore not easily visible on the plot

**Figure 2:**
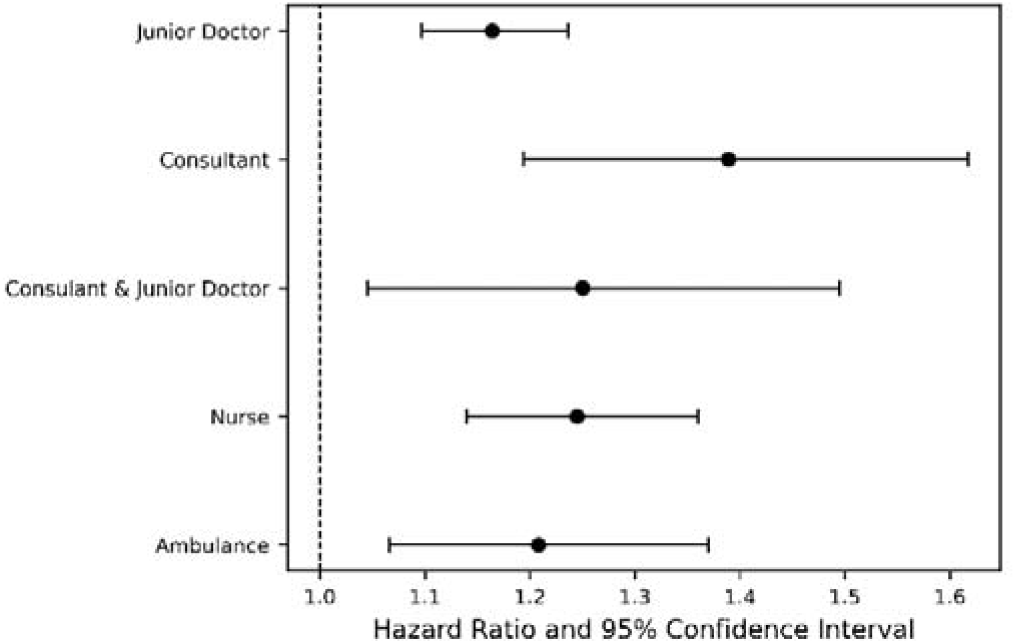
Forest plot demonstrating hazard ratios and confidence intervals of the impact of strikes on patients’ “time in ED” compared to the baseline of no strike – higher hazard implies patients move through the ED faster.

**Figure 3:**
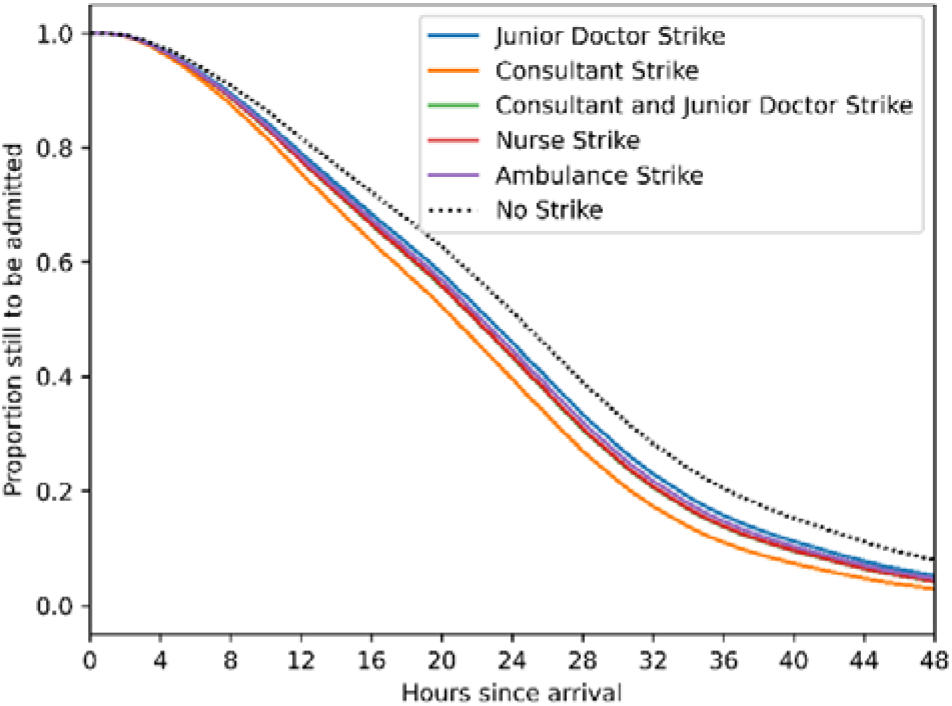
Fitted Cox-regression “time in ED” curves for each of the different strike types in the analysis.

### Limitations

The findings in this paper agree with other, international, literature on patient flow and ‘waiting time’ during the strikes. The impact of the strikes will likely vary by hospital. Furthermore, due to this paper’s focus on exit block, our analysis included only patients who were admitted to the hospital.

The times at which a patient is recorded to have arrived and been admitted are automatically logged when a healthcare worker updates the patient’s data. This manual choice can be seen in the spike in admissions immediately before the four-hour mark, which is likely due to healthcare workers attempting to meet targets. This control of recording by healthcare workers may mean changes in staff mix during the strikes may cause changes in recording.

The claims made about the similar results across all the strikes are based on the recorded striking group dates and assume that striking groups cover each other’s strikes equally. Employment of locum doctors, differences in staff volume, or certain sub-groups not striking during this time would not be accounted for and may skew results.

Our Cox-proportional hazards methodology has some limitations. The model falls short of completely satisfying the proportional hazards assumption. The assumptions were tested using Schoenfeld residuals and log-minus-log plots to assess validity of the results. However, it is well documented that the Cox-proportional hazards regression model is robust against such deviations from the model assumptions (20,30). Our hazard ratio estimates were calculated using a robust bootstrapping procedure to ensure maximum validity to our results. We believe that because of the robustness of the model and the model specification, our hazard ratios lie close to their true values.

### Future Work

Replicating this study in other settings would validate the results from this study. Future analyses should directly investigate the implied relationship between inpatient capacity and patient time in ED suggested from these results. Staffing data could be included in further analyses, fully accounting for staffing levels and staff seniority during the strike measures. Numbers and details of investigations taken may also give insight into the efficiency of the emergency department operations during the strikes.

Further investigation into the long-term effects of strikes should also be investigated. EDs could see an increase in attendances in the days after a strike due to postponement of those that would have attended during the strike. Postponement of elective activity may also result in emergency attendances in the longer-term due to conditions going untreated. To fully understand our results, we suggest a qualitative study to gain perspectives from patients, clinicians and other members of the NHS workforce affected by the strikes.

### Conclusion

There is a lack of studies on the effect of strikes on the healthcare system in general. The strikes provide an interesting counterfactual example to the status quo of NHS healthcare delivery. This paper is, to our knowledge, the first to investigate the effects of the recent NHS strikes on patient flow through the emergency department. The preparation for strike days – allowing additional inpatient capacity downstream, demonstrates that even despite the strike conditions flow through NHS emergency departments can be improved by additional capacity and efficient discharge of medically fit patients.

## Data Availability

Data supporting this study is not publicly available due to ethical considerations around accessing linked patient-level healthcare data. Please contact the main author for more information.

## Research Ethics Approval

The project was approved by Lancaster University Faculty of Health and Medicine Research Ethics committee, reference FHM-2023-3868.

