## Supplementary Material for "Evaluating the Impact of NHS Strikes on Patient Flow through Emergency Departments"

**Strike Dates**

Strike days that directly impacted the emergency department were investigated. Regional strikes outside of Lancashire and strikes for specific specialties were not accounted for. The online resource strikecalendar.co.uk was used to derive the dates. These dates were cross-referenced with a Wikipedia database of NHS strikes since the start of 2022, no discrepancies were found (24,25) a table of strike dates and associated striking groups used in the analysis, the table can be found in the supplementary materials.

Truth table for which strike dates are classed as which strike.

Table S 1: Strike dates included in the analysis with associated strike groups.

|  | Nurse strike | Ambulance strike | Junior doctor strike | Consultant strike |
| --- | --- | --- | --- | --- |
| 15/12/2022 | TRUE | FALSE | FALSE | FALSE |
| 20/12/2022 | TRUE | FALSE | FALSE | FALSE |
| 21/12/2022 | FALSE | TRUE | FALSE | FALSE |
| 11/01/2023 | FALSE | TRUE | FALSE | FALSE |
| 18/01/2023 | TRUE | FALSE | FALSE | FALSE |
| 19/01/2023 | TRUE | FALSE | FALSE | FALSE |
| 23/01/2023 | FALSE | TRUE | FALSE | FALSE |
| 06/02/2023 | TRUE | FALSE | FALSE | FALSE |
| 07/02/2023 | TRUE | FALSE | FALSE | FALSE |
| 10/02/2023 | FALSE | TRUE | FALSE | FALSE |
| 01/03/2023 | TRUE | FALSE | FALSE | FALSE |
| 02/03/2023 | TRUE | FALSE | FALSE | FALSE |
| 03/03/2023 | TRUE | FALSE | FALSE | FALSE |
| 13/03/2023 | FALSE | FALSE | TRUE | FALSE |
| 14/03/2023 | FALSE | FALSE | TRUE | FALSE |
| 15/03/2023 | FALSE | FALSE | TRUE | FALSE |
| 20/03/2023 | FALSE | TRUE | FALSE | FALSE |
| 11/04/2023 | FALSE | FALSE | TRUE | FALSE |
| 12/04/2023 | FALSE | FALSE | TRUE | FALSE |
| 13/04/2023 | FALSE | FALSE | TRUE | FALSE |
| 14/04/2023 | FALSE | FALSE | TRUE | FALSE |
| 15/04/2023 | FALSE | FALSE | TRUE | FALSE |
| 14/06/2023 | FALSE | FALSE | TRUE | FALSE |
| 15/06/2023 | FALSE | FALSE | TRUE | FALSE |
| 16/06/2023 | FALSE | FALSE | TRUE | FALSE |
| 13/07/2023 | FALSE | FALSE | TRUE | FALSE |
| 14/07/2023 | FALSE | FALSE | TRUE | FALSE |
| 15/07/2023 | FALSE | FALSE | TRUE | FALSE |
| 16/07/2023 | FALSE | FALSE | TRUE | FALSE |
| 17/07/2023 | FALSE | FALSE | TRUE | FALSE |
| 18/07/2023 | FALSE | FALSE | TRUE | FALSE |
| 20/07/2023 | FALSE | FALSE | FALSE | TRUE |
| 21/07/2023 | FALSE | FALSE | FALSE | TRUE |
| 22/07/2023 | FALSE | FALSE | FALSE | TRUE |
| 11/08/2023 | FALSE | FALSE | TRUE | FALSE |
| 12/08/2023 | FALSE | FALSE | TRUE | FALSE |
| 13/08/2023 | FALSE | FALSE | TRUE | FALSE |
| 14/08/2023 | FALSE | FALSE | TRUE | FALSE |
| 24/08/2023 | FALSE | FALSE | FALSE | TRUE |
| 25/08/2023 | FALSE | FALSE | FALSE | TRUE |
| 19/09/2023 | FALSE | FALSE | FALSE | TRUE |
| 20/09/2023 | FALSE | FALSE | TRUE | TRUE |
| 21/09/2023 | FALSE | FALSE | TRUE | FALSE |
| 22/09/2023 | FALSE | FALSE | TRUE | FALSE |
| 02/10/2023 | FALSE | FALSE | TRUE | TRUE |
| 03/10/2023 | FALSE | FALSE | TRUE | TRUE |
| 04/10/2023 | FALSE | FALSE | TRUE | TRUE |
| 20/10/2023 | FALSE | FALSE | TRUE | FALSE |
| 21/10/2023 | FALSE | FALSE | TRUE | FALSE |
| 22/10/2023 | FALSE | FALSE | TRUE | FALSE |
| 03/01/2024 | FALSE | FALSE | TRUE | FALSE |
| 04/01/2024 | FALSE | FALSE | TRUE | FALSE |
| 05/01/2024 | FALSE | FALSE | TRUE | FALSE |
| 06/01/2024 | FALSE | FALSE | TRUE | FALSE |
| 07/01/2024 | FALSE | FALSE | TRUE | FALSE |
| 08/01/2024 | FALSE | FALSE | TRUE | FALSE |
| 25/02/2024 | FALSE | FALSE | TRUE | FALSE |
| 26/02/2024 | FALSE | FALSE | TRUE | FALSE |
| 27/02/2024 | FALSE | FALSE | TRUE | FALSE |
| 28/02/2024 | FALSE | FALSE | TRUE | FALSE |
| 15/12/2022 | TRUE | FALSE | FALSE | FALSE |
| 20/12/2022 | TRUE | FALSE | FALSE | FALSE |
| 21/12/2022 | FALSE | TRUE | FALSE | FALSE |
| 11/01/2023 | FALSE | TRUE | FALSE | FALSE |

**Referral Destination Categories**

Referral destination categories in the ‘Other’ class are Geriatric, Cardiology, Nephrology, Obstetrics, Clinical oncology, Ophthalmology, Clinical oncology, Neurology, Rehabilitation, Endocrinology.

**Results**

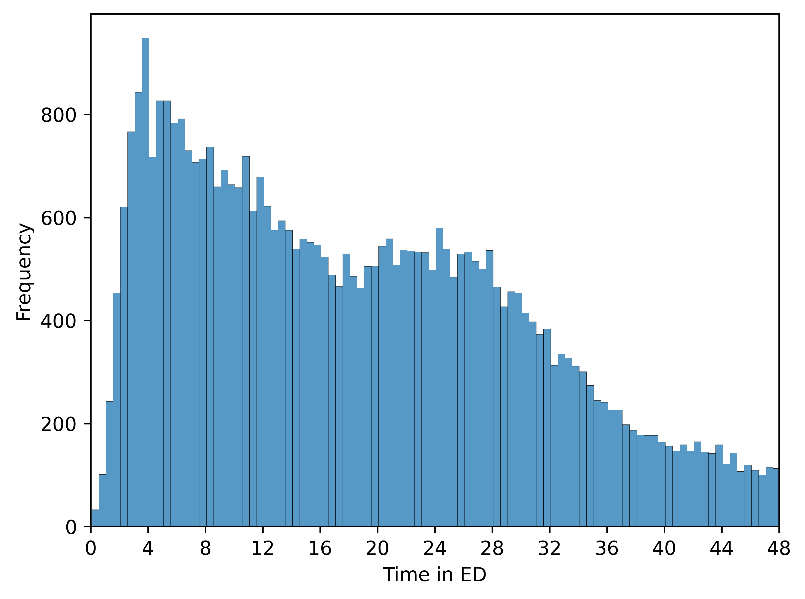

Figure S 1: Histogram of admitted patients' time spent in ED.

In Figure S 1 see a high, widespread distribution of patients’ “time in ED”. This is because we are looking at just patients who were admitted since the start of 2022. Admitted patients generally are in ED for longer, and patient “time in ED” has increased over time, so this is a subset of attendances with a high “time in ED”.

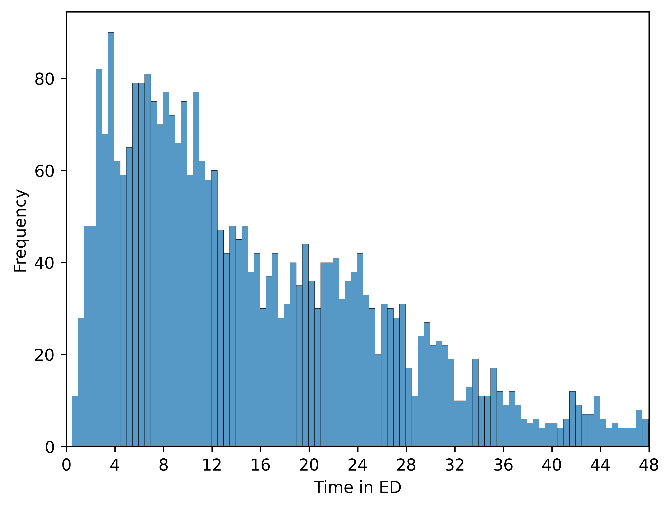
In Figure S 2, a similar distribution of “time in ED” can be seen for patients who are admitted on strike days.

Figure S 2: Histogram of admitted patients' time spent in ED on strike days.

**Kaplan Meier Exploratory Analysis**

We produced Kaplan-Meier plots to demonstrate “time in ED” curves of admitted patients, separated by categorical variables.

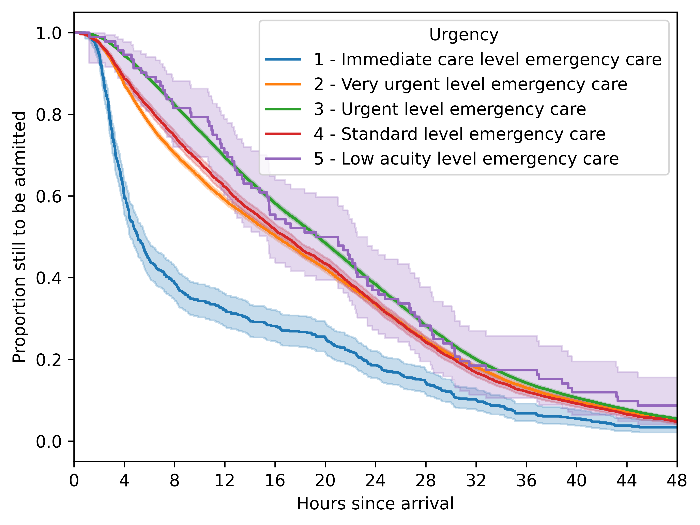

Figure S 3: Kaplan-Meier “time in ED” curves stratified by urgency.

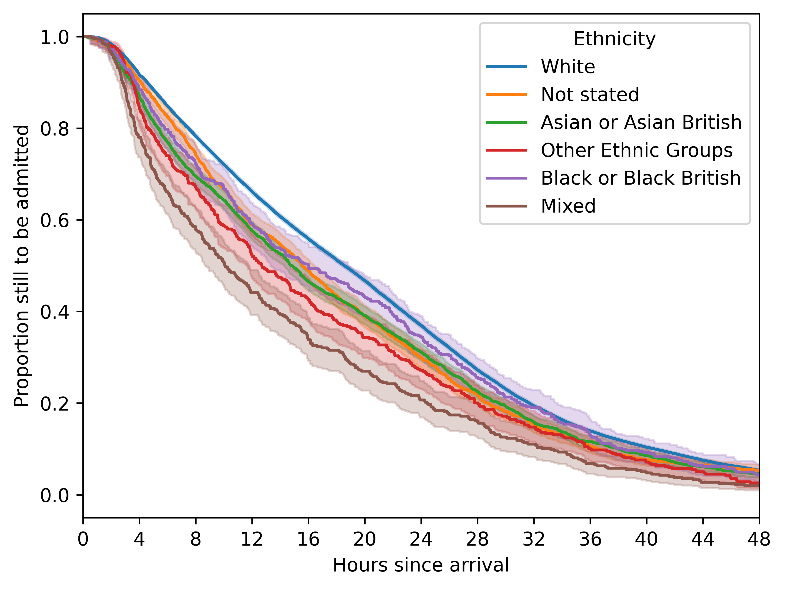

Figure S 4: Kaplan-Meier “time in ED” curves stratified by ethnicity.

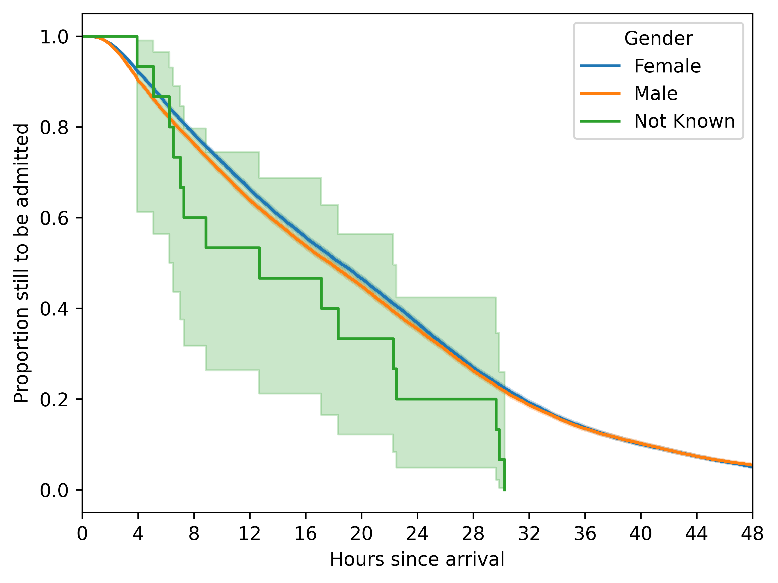

Figure S 5: Kaplan-Meier “time in ED” curves stratified by gender.

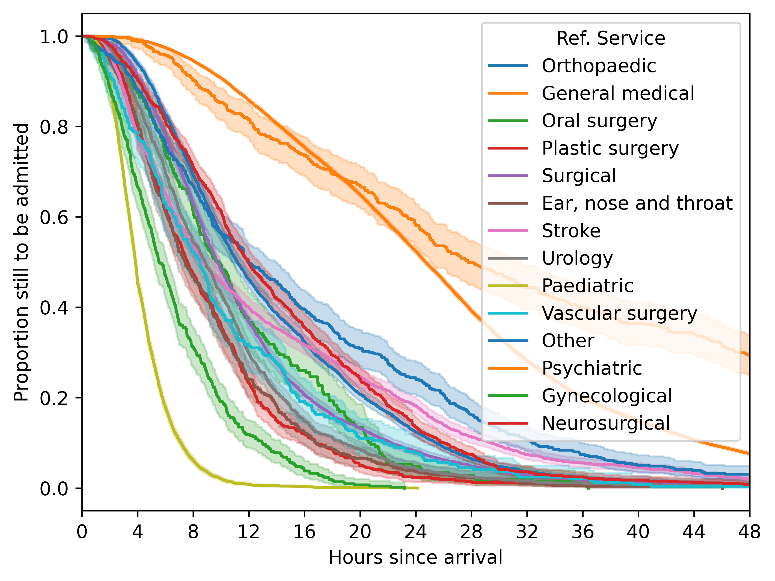

Figure S 6: Kaplan-Meier “time in ED” curves stratified by referral service.

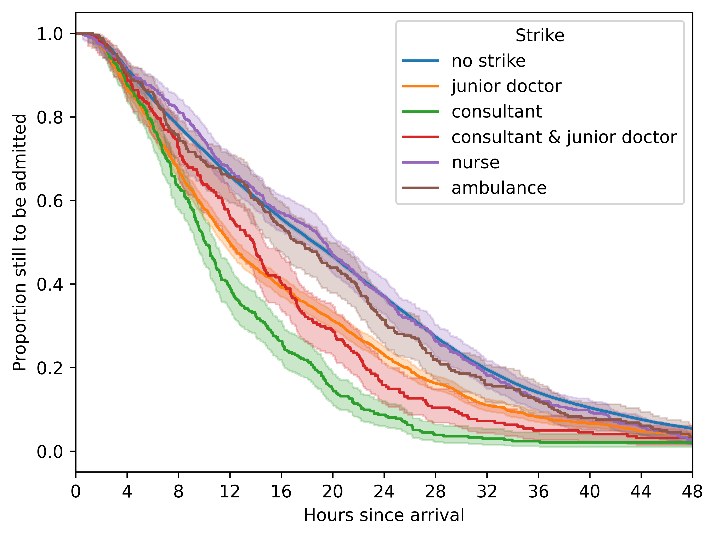

Figure S 7: Kaplan-Meier “time in ED” curves stratified by strike type.

**Tests for Proportional Hazards**

**Log-log plots**

We can assess the proportional hazards assumption by plotting the transformed survival (“time in ED”) function $S\left( t \right)$ against the log-transformed time. For the proportional hazards assumption to hold, the curves should be roughly parallel.

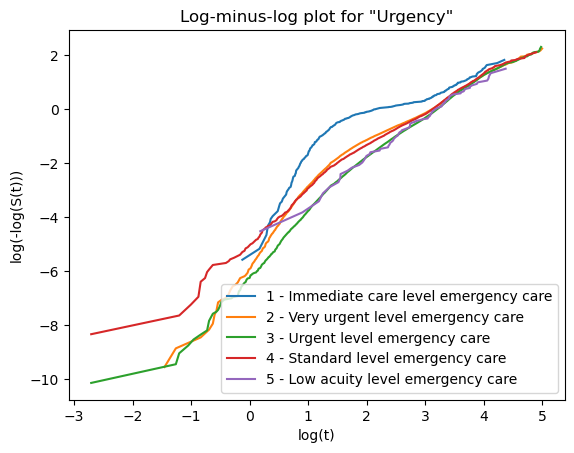

Figure S 8: Log-minus log plot of survival functions stratified by the Urgency variable.

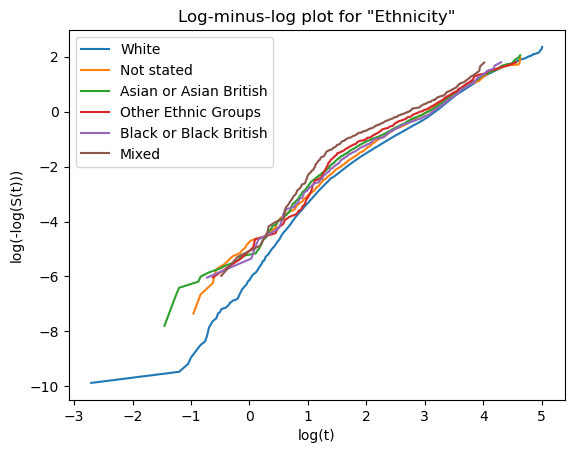

Figure S 9: Log-minus log plot of survival functions stratified by the Ethnicity variable.

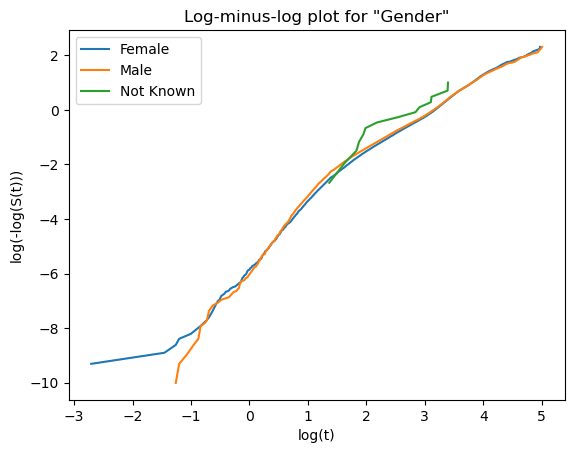

Figure S 10: Log-minus log plot of survival functions stratified by the Gender variable.

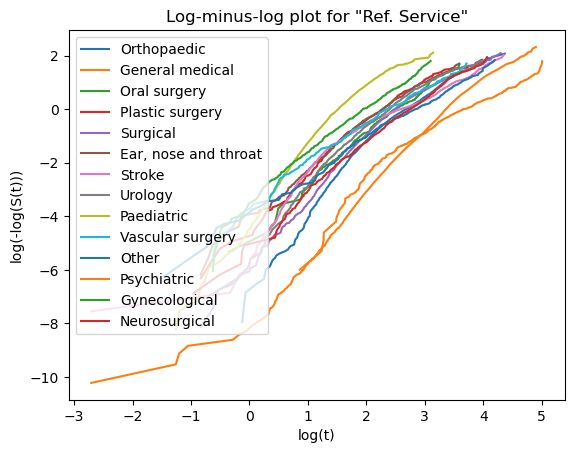

Figure S 11: Log-minus log plot of survival functions stratified by the Referral Service variable.

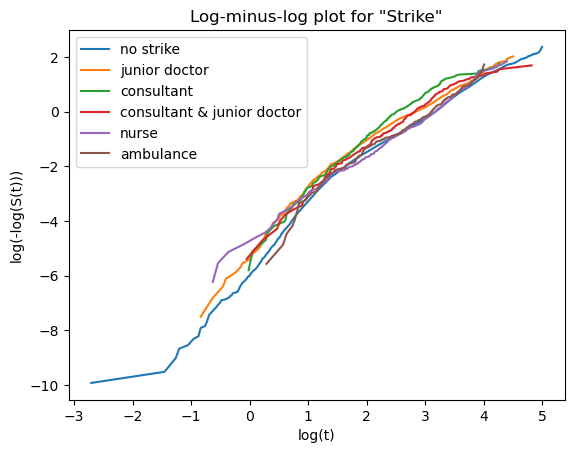

Figure S 12: Log-minus log plot of survival functions stratified by the Strike Type variable.

**Cox-Proportional Hazards**

Table S 2: Full coefficients, and corresponding hazards ratios, for the Cox-proportional hazards model.

| Variable | Level | Coefficient | Hazard Ratio | 95% CI | | p-value |
| --- | --- | --- | --- | --- | --- | --- |
| Linear time trend | - | 0.008 | 1.008 | 1.007 | 1.009 | 8.76E-37 |
| Yearly harmonic (sine) | - | 0.018 | 1.018 | 1.004 | 1.032 | 0.009077 |
| Yearly harmonic term (cosine) | - | -0.073 | 0.93 | 0.916 | 0.943 | 2.02E-22 |
| Daily harmonic term (sine) | - | -0.011 | 0.989 | 0.976 | 1.002 | 0.092833 |
| Daily harmonic term (cosine) | - | 0.008 | 1.008 | 0.995 | 1.021 | 0.236537 |
| ED heat | - | -0.013 | 0.987 | 0.987 | 0.988 | 0 |
| Urgency | 2 | -0.41 | 0.664 | 0.608 | 0.725 | 3.91E-20 |
| Urgency | 3 | -0.456 | 0.634 | 0.581 | 0.691 | 8.72E-25 |
| Urgency | 4 | -0.395 | 0.674 | 0.615 | 0.738 | 1.73E-17 |
| Urgency | 5 – least urgent | -0.49 | 0.613 | 0.491 | 0.765 | 1.51E-05 |
| Referral Service | General medical | -1.104 | 0.331 | 0.319 | 0.345 | 0 |
| Referral Service | Oral surgery | 0.204 | 1.226 | 1.064 | 1.413 | 0.004788 |
| Referral Service | Plastic surgery | 0.505 | 1.657 | 1.512 | 1.817 | 4.92E-27 |
| Referral Service | Surgical | 0.164 | 1.179 | 1.121 | 1.239 | 1.21E-10 |
| Referral Service | Ear, nose and throat | 0.526 | 1.692 | 1.535 | 1.864 | 2.40E-26 |
| Referral Service | Stroke | -0.144 | 0.866 | 0.817 | 0.918 | 1.44E-06 |
| Referral Service | Urology | 0.444 | 1.56 | 1.453 | 1.674 | 5.23E-35 |
| Referral Service | Pediatric | 1.763 | 5.83 | 5.487 | 6.195 | 0 |
| Referral Service | Vascular surgery | 0.412 | 1.509 | 1.324 | 1.72 | 6.79E-10 |
| Referral Service | Other | -0.377 | 0.686 | 0.624 | 0.754 | 4.29E-15 |
| Referral Service | Psychiatric | -2.197 | 0.111 | 0.1 | 0.124 | 0 |
| Referral Service | Gynaecology | 1.09 | 2.974 | 2.679 | 3.302 | 9.79E-93 |
| Referral Service | Neurosurgical | -0.153 | 0.858 | 0.798 | 0.924 | 4.50E-05 |
| Age |  | -0.008 | 0.992 | 0.992 | 0.993 | 1.05E-208 |
| Ethnicity | Not Stated | -0.016 | 0.984 | 0.935 | 1.035 | 0.529732 |
| Ethnicity | Asian or Asian British | 0.015 | 1.015 | 0.974 | 1.058 | 0.488383 |
| Ethnicity | Other Ethnic Groups | -0.057 | 0.945 | 0.857 | 1.041 | 0.247738 |
| Ethnicity | Black or Black British | 0.011 | 1.011 | 0.919 | 1.113 | 0.818126 |
| Ethnicity | Mixed | 0.049 | 1.05 | 0.95 | 1.16 | 0.338741 |
| Gender | Male | -0.011 | 0.989 | 0.97 | 1.008 | 0.244186 |
| Gender | Not known | 0.39 | 1.477 | 0.89 | 2.451 | 0.131611 |
| Strike type | Junior doctor | 0.224 | 1.251 | 1.189 | 1.316 | 3.19E-18 |
| Strike type | Consultant | 0.333 | 1.396 | 1.25 | 1.558 | 3.02E-09 |
| Strike type | Consultant & junior doctor | 0.236 | 1.266 | 1.107 | 1.447 | 0.000569 |
| Strike type | Nurse | 0.221 | 1.247 | 1.142 | 1.362 | 9.32E-07 |
| Strike type | Ambulance | 0.191 | 1.211 | 1.072 | 1.368 | 0.002049 |

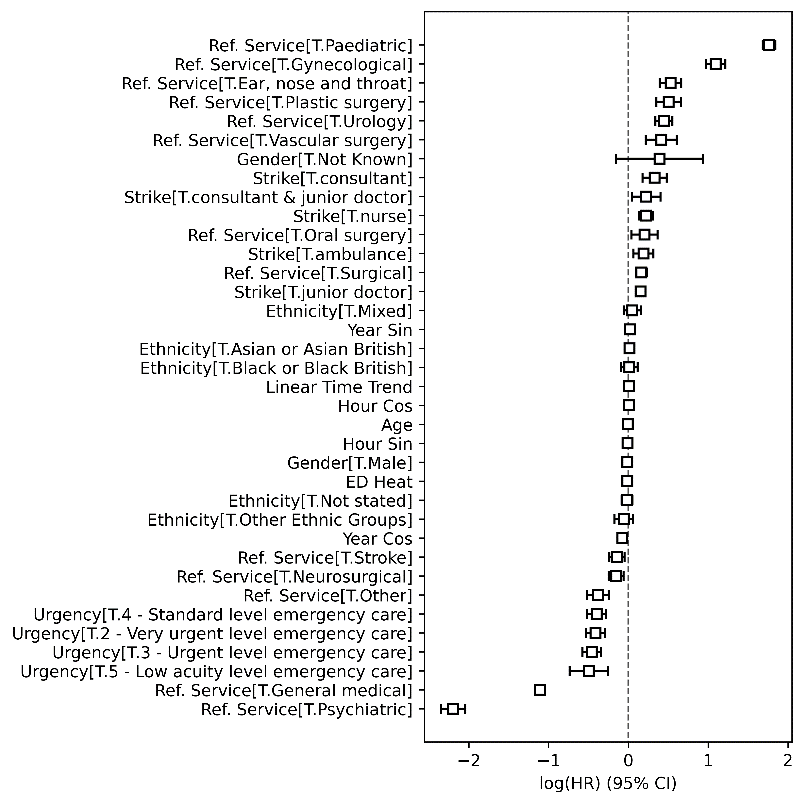

Figure S 13: Forrest plot for all variables included in the model.

### ED2 Analysis

**Exploratory Kaplan-Meier Curves**

The model for ED did not include referral service, due to low variance between categories and therefore convergence issues. This similarity between referral service “time in ED” curves can be seen in the Kaplan-Meier plot in Figure S 14.

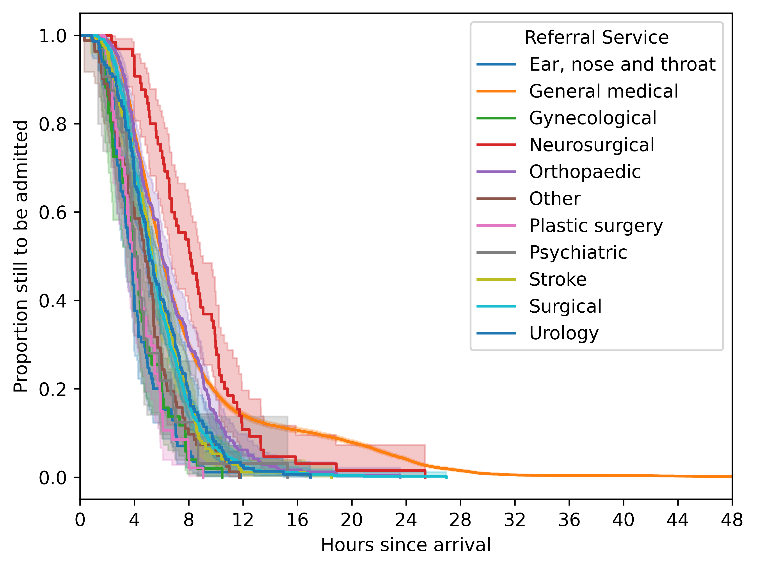

Figure S 14: ED2 - Kaplan-Meier curves of admitted patients' “time in ED”, separated by referral service they are admitted to. The shaded area around each line represents the 95% confidence interval (CI) for the KM estimate at each point.

**Cox-Proportional Hazards Model**

The coefficients and hazard ratios for the strikes at ED2 are shown in Table S 3. The results from ED2 are less consistent. Most of the strikes appear to have no significant impact on flow. The only significant hazard ratio is for consultant strikes, with a hazard ratio of 1.36. The entire model coefficients and hazard ratios can be found in Table S 4.

Table S 3:ED2 - Coefficients and hazard ratios for the strike variables in the Cox-proportional hazards model. Here, higher hazard refers to a higher likelihood of being admitted into the hospital.

| Strike Type | Coefficient | Hazard Ratio | 95% CI | | p-value | |
| --- | --- | --- | --- | --- | --- | --- |
| Baseline – No strike |  |  | - | - | | - |
| Junior doctor strike | -0.075 | 0.927 | 0.844 | 1.02 | | 0.119217 |
| Consultant strike | 0.306 | 1.358 | 1.098 | 1.679 | | 0.004686 |
| Consultant and junior doctor strike | -0.264 | 0.768 | 0.599 | 0.985 | | 0.037539 |
| Nurse strike | -0.03 | 0.97 | 0.827 | 1.138 | | 0.709028 |
| Ambulance strike | 0.283 | 1.327 | 1.065 | 1.653 | | 0.011718 |
| ** Statistically significant* | | | | | | |

The resulting fitted “time in ED” curves from the Cox model are shown in Figure S 15. In this case it is visible that “time in ED” reduces for some strikes and increases for others.

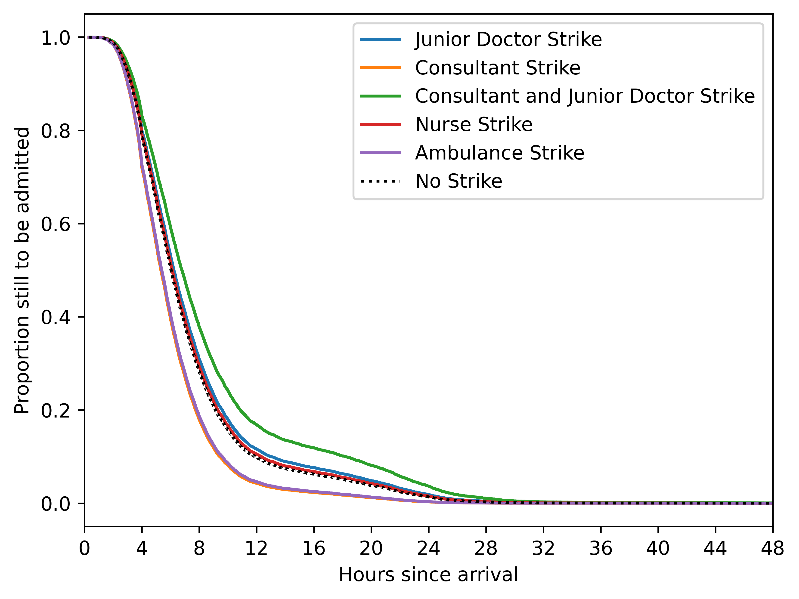

Figure S 15: ED2 - Fitted Cox-regression “time in ED” curves for each of the different strike types in the analysis.

Full model coefficients for ED2 can be found in

Table S 4: ED2 – Full coefficients, and corresponding hazards ratios, for the Cox-proportional hazards model.

| Variable | Level | Coefficient | Hazard Ratio | 95% CI | | p-value |
| --- | --- | --- | --- | --- | --- | --- |
| Linear time trend |  | -0.039 | 0.962 | 0.959 | 0.964 | 1.40E-157 |
| Yearly harmonic (sine) |  | -0.05 | 0.951 | 0.924 | 0.979 | 0.00075 |
| Yearly harmonic term (cosine) |  | -0.189 | 0.828 | 0.805 | 0.852 | 3.41E-39 |
| ED heat |  | -0.022 | 0.978 | 0.976 | 0.98 | 2.20E-94 |
| Urgency | 2 | 0.009 | 1.009 | 0.676 | 1.506 | 0.965756 |
| Urgency | 3 | -0.089 | 0.915 | 0.614 | 1.365 | 0.664153 |
| Urgency | 4 | -0.118 | 0.889 | 0.595 | 1.328 | 0.566124 |
| Urgency | 5 – least urgent | 0.041 | 1.042 | 0.524 | 2.071 | 0.906592 |
| Age |  | -0.003 | 0.997 | 0.995 | 0.998 | 4.73E-10 |
| Ethnicity | Not Stated | -0.09 | 0.914 | 0.818 | 1.l022 | 0.115352 |
| Ethnicity | Asian or Asian British | 0.115 | 1.122 | 0.921 | 1.367 | 0.253371 |
| Ethnicity | Other Ethnic Groups | -0.099 | 0.906 | 0.687 | 1.195 | 0.483723 |
| Ethnicity | Black or Black British | -0.154 | 0.857 | 0.605 | 1.215 | 0.387172 |
| Ethnicity | Mixed | -0.231 | 0.793 | 0.517 | 1.218 | 0.289622 |
| Gender | Male | 0.051 | 1.052 | 1.013 | 1.093 | 0.009273 |
| Gender | Not known | -0.035 | 0.966 | 0.475 | 1.964 | 0.923696 |
| Strike type | Junior doctor | -0.075 | 0.927 | 0.844 | 1.02 | 0.119217 |
| Strike type | Consultant | 0.306 | 1.358 | 1.098 | 1.679 | 0.004686 |
| Strike type | Consultant & junior doctor | -0.264 | 0.768 | 0.599 | 0.985 | 0.037539 |
| Strike type | Nurse | -0.03 | 0.97 | 0.827 | 1.138 | 0.709028 |
| Strike type | Ambulance | 0.283 | 1.327 | 1.065 | 1.653 | 0.011718 |
